# Determinants of stunting among children < 5 years of age: Evidence from 2018-2019 Zambia Demographic and Health Survey

**DOI:** 10.1101/2021.05.19.21257389

**Authors:** Bright Nkhoma, Wingston Felix Ng’ambi, Peter J. Chipimo, Mowa Zambwe

## Abstract

**OBJECTIVE:** The study aimed at identifying socio-economic and environmental factors that were associated with stunting among children aged 0-59 months in Zambia.

**BACKGOUND:** Hitherto, stunting continues to be a Public Health problem worldwide. A child is ‘stunted’ if his or her height is less than negative two standard deviations below the World Health Organization (WHO) standard. The study aimed to explore determinants of stunting in Zambia among children (< 5 years of age) using the Zambia Demographic and Health Survey (ZDHS) 2018-2019 database.

**METHODS:** A total of 7, 045 Zambian children with complete anthropometric measurements and aged 0–59 months were included in the study. Nutritional status was evaluated using anthropometric; height-for-age, as a proxy measure of stunting. Univariate and multivariate binary logistic regression were used to examine the association between stunting and selected environmental, maternal-socio-demographic and child level variables.

**RESULTS:** A total of 2, 479 children under the age of five found to be stunted representing a prevalence of 34.9%. Stunting was higher among male children as compared to female children (38.5% vs 31.3% respectively). Additional analysis revealed that children from households whose source of drinking water was improved (34%) were less likely to be stunted compared to children from households whose source of drinking water was non-improved (40%). Stunting was statistically significant associated with sex and age of a child; birth size; breastfeeding; residence; maternal education; wealth index; twin births and the birth interval among siblings. Children born to mothers whose previous birth interval is less than 24 months (aOR= 1.34 95%CI: 1.13-1.58; p<0.001), children from lower index households (aOR= 1.65 95%CI: 1.32-2.08; p<0.001), twin births (aOR=2.65 95%CI: 1.61-4.36; p<0.001), children whose mothers had primary education (aOR=1.16 95%CI 1.00-1.35; p=0.046), children coming from households whose source of drinking water was non-improved (aOR= 1.30 95%CI: 1.09-1.5: p=0.003), child not breastfed (aOR= 1.20 95%CI: 1.04-1.38; p=0.015) were more likely to be stunted.

**CONCLUSION:** The study established that the major predictors of stunting among children under 5 years old in Zambia were sex and age of the child; birth weight; maternal education; wealth status; source of drinking water; twin births, breastfeeding, residence and the birth interval among siblings. Therefore, to reduce the burden of stunting interventions that can address these factors are required such as community based education and targeted nutritional interventions.

**ARTICLE SUMMARY:** *Strengths and limitations of the study:*

1. The study utilised the ZDHS data as a proxy to establish the national burden of stunting in Zambia during 2018/2019, hence contributing to the body of knowledge.
2. Many variables perataining to food intakes were not covered in ZDHS of 2018/2019 hence exluded from the study. Food intakes are very critical in measuring stunting levels.

## Introduction

Hitherto, stunting or low Height-for-Age z-score (HAZ) continues to be a Public Health problem affecting growth potential in children. Child stunting refers to a child who is too short for his or her age and is the result of chronic or recurrent malnutrition. A child is ‘stunted’ if his or her height is less than negative two standard deviations below the World Health Organization standard [17].

Stunting is a contributing risk factor to child mortality and is also a marker of inequalities in human development. Stunted children fail to reach their physical and cognitive potential. Stunting is more prevalent in developing countries such as many countries in Asia and Africa stunting. According to the United Nations, 35.6% and 29.1% of children in Eastern and Southern Africa, under 5 years of age are stunted, respectively [11, 18]. Over the past years, the prevalence of stunting has gradually reduced but generally progress is unsatisfactory as countless children are at risk. At national level stunting prevalence has persistently remained above 35% among children below the age of 5 with Northern Province recording the highest prevalence at 46% [19].

Stunted growth is as the result of several prevailing factors vis-à-vis; poor maternal health and nutrition, inadequate infant and young child feeding practices, infection, health, water and sanitation services, demographic and socio-economic factors. In terms of maternal health and nutrition, these include: maternal nutritional and health status before, during and after pregnancy that influences a child’s early growth and development, commencement in the womb. Infant and young child feeding practices that contribute to stunting include suboptimal breastfeeding (specifically, non-exclusive breastfeeding) and balancing feeding that is limited in quantity, quality and variety [10–15].

Stunting is a huge drain on economic productivity and development because resources that can be prorated to other infrastructural development are altered to nutrition and feeding programs, and sensitization of mothers on good feeding practices. Economists estimate that stunting can reduce a country’s Gross Domestic Product (GDP) by up to 3% [7]. More so, stunting leads to short adult height, long-term effects on individuals and societies; including diminished cognitive and physical development, reduced productive capacity and poor health, and an increased risk of degenerative diseases such as diabetes.

In Zambia, evidence regarding factors associated with stunting has been very low and limited. So, there is need to continually analyse the factors anchoring stunting in Zambia as the way of finding new causes.

Stunting has a number of effects on children, which later shows into adulthood and negatively impact the nation as a whole. The resulting findings would inform policy and programmes and relevant in decision making and knowledge generation. Thus, the purpose of this study was to identify socio-economic and environmental factors that were associated with stunting among children aged 0-59 months in Zambia.

## Materials and Methods

The study extracted data for children (< 5 years of age) from the 2018/2019 Zambia Demographic Health Survey database. The dataset provided data on child anthropometric measurements, socio-economic variables, food types and other variables. These variables were selected based on United Nations Children’s Emergency Fund (UNICEF) framework of the factors that determine nutritional status [16]. The survey was a cross sectional which was a nationally representative probability sample of women in the reproductive age 15 to 49 and men 15 to 59 years.

### Sampling and data collection methods

The 2018 ZDHS followed a stratified two-stage sample design. The first stage involved selecting sample points (clusters) consisting of EAs. EAs were selected with a probability proportional to their size within each sampling stratum. A total of 545 clusters were selected. The second stage involved systematic sampling of households. A household listing operation was undertaken in all of the selected clusters. During the listing, an average of 133 households were found in each cluster, from which a fixed number of 25 households were selected through an equal probability systematic selection process, to obtain a total sample size of 13,625 households. Results from this sample are representative at the national, urban and rural, and provincial levels. All women aged 15-49 and men aged 15-59 who were either permanent residents of the selected households or visitors who stayed in the households the night before the survey were eligible to be interviewed.

### Statistical Analysis

The main objective of the study was to identify factors that were associated with stunting in Zambia. The ZDHS dataset was de-identified to prevent a person’s identity from being linked with information. First, a binary variable was created defining stunting as “stunted” (Z-score less than −2 SD) and “not stunted” (Z-score equal to and greater than −2 SD). Based on literature and guidance from the conceptual framework the study included the following independent variables: child sex, child age, immunisation, antenatal care during pregnancy, place of delivery, maternal education, age of mother, mother’s previous birth interval, birth size, current marital status, twins births, birth intervals among siblings, number of children ever born, breastfeeding, residence, wealth index, and quality of source of drinking water. For the quality of source of drinking water, a variable was created categorising the sources as either being improved (piped water, protected well and spring, bottled water and rainwater) and non-improved (unprotected well and spring, tanker, surface water and other). Statistical analysis was performed using Stata version 14 (StataCorp, College Station, Texas, USA). With relatively large sample, the distribution of data was assumed to be normal meaning it is representative of the population. Pearson’s Chi-Square test (measure of relationship between two categorical variables) was used to explore relationships between prevalence of stunting and the independent variables. Reported p<0.05 and 95% CI was considered statistically significant meaning there is an association between independent variable and stunting. Since the outcome variable is binary (“Stunted” or “Not Stunted”) multiple logistic regression was done to measure the net associations of all variables on stunting. Only predictors with significant p-values (that is p < 0.05 and 95% CI) was considered for multiple logistic analysis. Except for maternal age, marital status, residence and children ever born, these were identified as important variable hence included in the model despite statistically insignificant at bivariate level. In the multiple regression, the final model was generated using a researcher led forward stepwise regression. Variables entered in the model and associations were considered statistically significant when p<0.05. The Odds ratios determined the odds of being stunted or not stunted.

## Results

### Proportion of respondents by demographic and socio-economic characteristics

**Table 1** shows the distribution of respondents by background characteristics. The majority were in the age range 30-39 (34%), 32.4% were in the age range 15-24, 25.7% were in the age range 25-29 and the minority were in the age range 40-49 (8%).

**Table 1:** Proportion of respondents by demographic and socio-economic characteristics

| Background Characteristics | Number | Percent |
| --- | --- | --- |
| **Total** | **7,045** | **100.0** |
| **Maternal Age** |  |  |
| 15-24 | 2283 | 32.4 |
| 25-29 | 1809 | 25.7 |
| 30-39 | 2393 | 34.0 |
| 40-49 | 560 | 8.0 |
| **Residence** |  |  |
| Urban | 1950 | 32.6 |
| Rural | 5095 | 67.4 |
| **Child Age in Months** |  |  |
| <6 | 773 | 10.9 |
| 6-11 | 717 | 10.1 |
| 12-23 | 1499 | 21.1 |
| Overall | 4056 | 57.8 |
| **Child Sex** |  |  |
| Male | 3477 | 49.4 |
| Female | 3568 | 50.6 |
| **Region** |  |  |
| Central | 648 | 8.0 |
| Copperbelt | 645 | 12.2 |
| Eastern | 875 | 14.2 |
| Luapula | 816 | 9.4 |
| Lusaka | 738 | 14.8 |
| Muchinga | 643 | 6.3 |
| Northern | 757 | 9.8 |
| North western | 579 | 5.6 |
| Southern | 729 | 12.9 |
| Western | 615 | 6.9 |
| **Education** |  |  |
| No education | 788 | 10.9 |
| Primary | 3778 | 52.4 |
| Secondary and Higher | 2479 | 36.7 |
| **Wealth Index** |  |  |
| Poor | 3917 | 50.8 |
| Middle | 1366 | 18.2 |
| Rich | 1762 | 31.0 |
| **Marital Status** |  |  |
| Ever Married | 6487 | 92.1 |
| Never Married | 558 | 7.9 |

In terms of residence, 67.4% resided in the rural area whereas as 34.6% in urban area. Nearly 58% of the children were aged 24-59 months, 21.1% were aged 12-23 months, 10.9% were aged less than 6 months and the least were aged 6-9 (10.1%). In terms of child sex, 50.6% were female and 49.4 were male.

In terms of region, 14.8% were from Lusaka, 14.2% from Eastern, 12.9% from Southern, 12.2% from Copperbelt, 9.8% from Northern, 9.4% from Luapula, 8% were Central, 6.9% from Western, 6.3% from Muchinga and 5.6 from North western.

The majority of these women, 52.4% had attained at least primary education (1-7th grade), 36.7% attained secondary (8-12th grade) and tertiary, 10.9% had no education. Slightly above half (50.8%) were in the low wealth index, 31% from high wealth index and the lowest 18.2% were from medium wealth index.

Over 90% of the participants reported being married (separated, divorced, married and widowed) while 7.9% reported as being never married.

### Prevalence of stunting in children under five by characteristics of children

**Table 2** shows that, 41.7% of children aged 12-23 months are stunted (p<0.001). Prevalence increased with increase in age. Disaggregation of this data by sex shows that male children are more stunted (38.5%) compared to female children (31.3%) (p<0.001). First born children are less likely to be stunted compared to children who are born as 1st multiple and 2nd multiple twins: 38.8% and 44.2% respectively (p<0.001).

**Table 2:** Prevalence of stunting in children under five by characteristics of children

| **Child Factors** | **Number** | **Stunted (%)** | **P-value** |
| --- | --- | --- | --- |
| Total | 2,479 | 34.9 |  |
| **Age in Months** |  |  |  |
| <6 | 162 | 18.6 |  |
| 6-11 | 185 | 25 | <0.001 |
| 12-23 | 619 | 41.7 |  |
| 24-59 | 1513 | 37.2 |  |
| **Sex of children** |  |  |  |
| Male | 1335 | 38.5 | <0.001 |
| Female | 1144 | 31.3 |  |
| **Twin Births** |  |  |  |
| Singe Birth | 2370 | 34.2 |  |
| 1st of multiple | 54 | 38.8 | <0.001 |
| 2nd of multiple | 55 | 44.2 |  |
| **Child Size** |  |  |  |
| Large | 504 | 29.2 |  |
| Average | 1525 | 34.7 | <0.001 |
| Small | 375 | 47.8 |  |
| **Immunisation** |  |  |  |
| Fully Immunised | 2030 | 34.7 | 0.692 |
| Not Immunised | 449 | 35.5 |  |

Further, stunting increases with decreasing reported size at birth: 47.8% of children born small (very small/smaller than average) are stunted compared to those born with average and large size (very large/larger than average) 45.9% and 34.7%% respectively (p<0.001). Those children not immunized are slightly likely to be stunted compared to children fully immunized (35.5% and 34.7 respectively: p=0.0692).

### Prevalence of stunting in children under five by characteristics of mother

**Table** 3 shows that stunting in children was high among mother’s aged 15–24 (36.4%: p=0.446). Children in rural areas were likely to be more stunted (35.4%) compared to children in urban areas (33.7%; p=0.282). Disaggregation by province shows that, Northern had the highest prevalence of children stunted 45.5% followed by Luapula 44.8% (p<0.001).

**Table 3:** Prevalence of stunting in children under five by characteristics of mother

| Maternal Factors | Number | Stunted (%) | P-value |
| --- | --- | --- | --- |
| Total | 2,479 | 34.9 |  |
| Age |  |  |  |
| 15-24 | 826 | 36.4 | $p=0.446$ |
| 25-29 | 632 | 34.5 |  |
| 30-39 | 823 | 33.9 |  |
| 40-49 | 198 | 34.2 |  |
| **Residence** |  |  |  |
| Urban | 654 | 33.7 | $p=0.282$ |
| Rural | 1825 | 35.4 |  |
| Province |  |  |  |
| Central | 220 | 34.0 | $p<0.001$ |
| Copperbelt | 208 | 29.9 |  |
| Eastern | 291 | 32.2 |  |
| Luapula | 372 | 44.8 |  |
| Lusaka | 264 | 37.6 |  |
| Muchinga | 206 | 32.5 |  |
| Northern | 339 | 45.5 |  |
| North western | 198 | 32.8 |  |
| Southern | 211 | 30.4 |  |
| Western | 170 | 27.5 |  |
| **Maternal Education** |  |  |  |
| No Education | 318 | 38.6 | $p=0.003$ |
| Primary | 1404 | 36.5 |  |
| Secondary and Higher | 757 | 31.4 |  |
| **Wealth Index** |  |  |  |
| Poor | 1509 | 37.9 | p<0.001 |
| Middle | 452 | 32.8 |  |
| Rich | 518 | 31.1 |  |
| Marital Status |  |  |  |
| Never Married | 204 | 35.9 | p=0.653 |
| Ever married | 2275 | 34.8 |  |

**Table 4:**
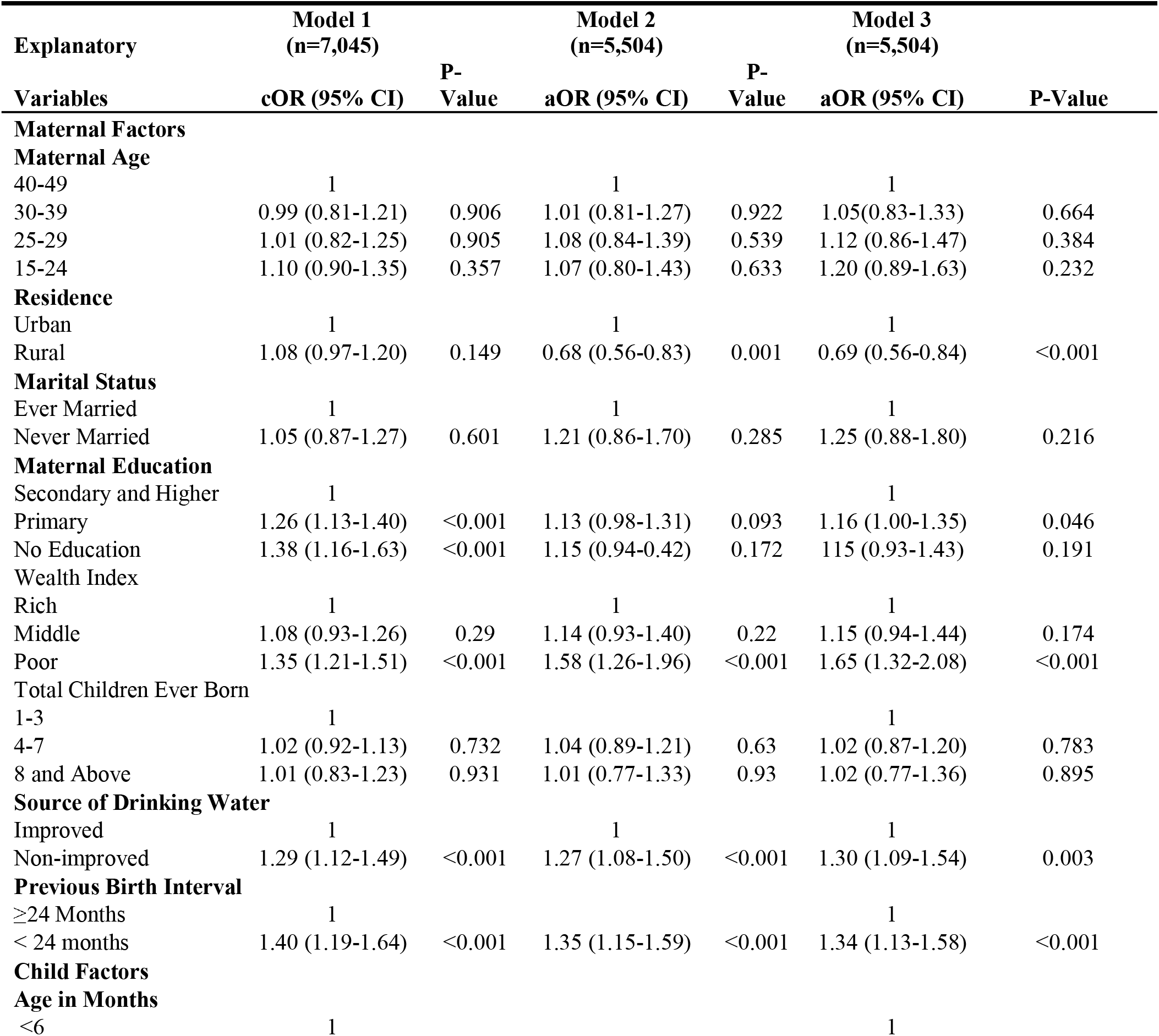
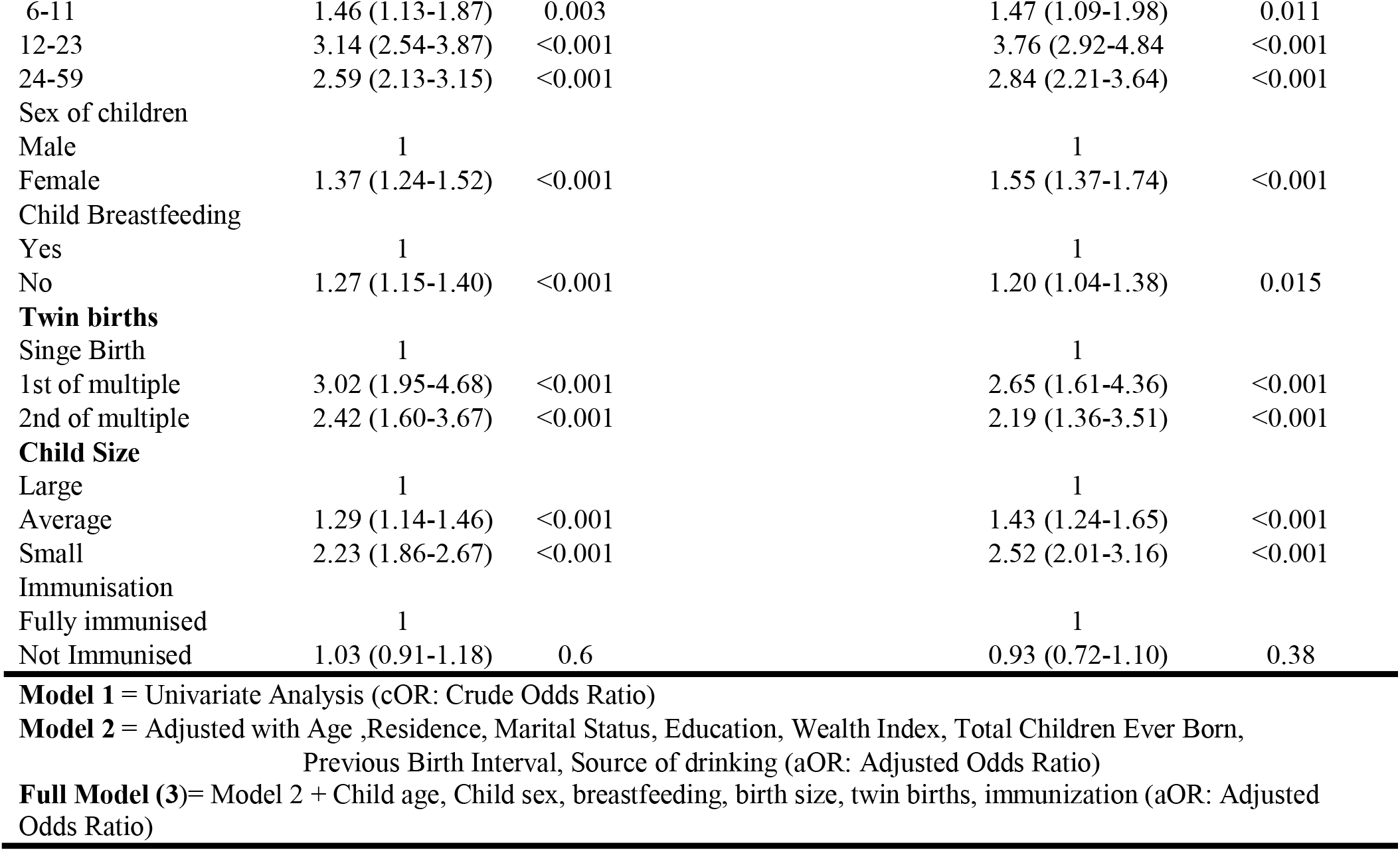
Univariate and multivariate analysis of Stunting (Height-for-age index)

Children whose mothers have attained secondary and higher education are less likely to be stunted 31.4% compared with children whose mothers had less education; (no education 38.6% and primary 32.9%; p<0.001). In the same way, children in households with wealth index described as poor are more likely to be stunted (37.9%) compared to those whose households are or were classified as belonging to the richer quintile 31.1% or middle (32.8%; p<0.001).

Stunting was slightly higher among children whose mother reported their marital status as “never married” (35.9%) compared to those mother’s reporting ever married (34.8%), although this was not statically significant (p=0.653).

By parity, stunting was more slightly pronounced in mother’s reporting to have had 4-7 children ever born (35.1%) than those with 8 and above children 34.9% and those with 1-3 children 34.7%: p=0.953). The findings further shows that, children whose mothers delivered at home were likely to be stunted (37.6%) than those whose mothers delivered at a healthy facility (34.3%: p=0.173).

Children from households whose source of drinking water was improved (34.0%) were less likely to be stunted compared to children from households whose source of drinking water was poor (40%: p=0.005) and this is attributed to the fact that non-improved water sources may be contaminated which may increase risk of infection such as diarrhoea.

Among the children who are stunted, 32.2% were currently being breastfed during the time of the survey compared with 37.6% who were not (p<0.001). Children whose mothers did not go for antenatal (40.9%) were likely to be stunted compared to children whose mothers went for antenatal less than four visits, and four and more visits (33.9% and 34.7% respectively: p=0.307).

About 41.5% of the children whose mothers previous birth interval is less than 24 months are stunted compared with 33.7% whose mothers previous birth interval is 24 or more (p<0.001).

### Factors Associated with Stunting Among Children

In binary logistic regression Adjusted Odds Ratios (AORs) residence, wealth index, maternal education, source of drinking water, previous birth interval were identified to be maternal factors associated with stunting. Furthermore, age in months, child, sex, breastfeeding, twin births, size at birth were identified as factors associated with stunting among children aged 0–59 months.

Univariate analysis (Model 1) indicated that children born to mothers who had no education (cOR= 1.38, 95%CI: 1.16-1.63), children in households with wealth quintiles described as poor (cOR= 1.35, 95%CI: 1.21-1.51), children in households whose source of drinking water classified as non-improved (cOR= 1.29, 95%CI: 1.12-1.49), mothers previous birth interval is less than 24 months (cOR= 1.40 95%CI: 1.19-1.64) were more likely to be stunted. In term of child factors, child aged 24-59 months (cOR= 2.59 95%CI: 2.13-3.15), female children (cOR= 1.37 95%CI: 1.24-1.52), child not breast during the time of the survey (cOR= 1.27 95%CI: 1.15-1.40), children born as 2nd multiple twins (cOR= 2.42 95%CI: 1.60-3.67) and size of the child at birth classified as small (cOR= 2.23 95%CI: 1.86-2.67) were more likely to be stunted.

In full model (aOR), children whose mothers resided in rural area (aOR= 0.69 95%CI: 0.56-0.84: p<0.001) were less likely to be stunted than those whose mothers resided in urban. Children whose mothers had primary education were more likely to be stunted compared with those whose mothers had secondary and higher education (aOR=1.16 95%CI 1.00-1.35; p=0.046).

Children (aged 0–59 months) from poor households (aOR= 1.65 95%CI: 1.32-2.08; p<0.001) were more likely to be stunted compared with those from richer households. Children coming from households whose source of drinking water was classified as non-improved (aOR= 1.30 95%CI: 1.09-1.5: p=0.003) were more likely to stunted than those coming from household whose source of drinking water is improved. Children born to mothers whose previous birth interval is less than 24 months (aOR= 1.34 95%CI: 1.13-1.58; p<0.001) more likely to be stunted than those whose mother previous birth interval is 24 months or more.

Age of the child is also associated with stunting. As age increases, there is an increase in the odds of a child being stunted (aOR= 3.76 95%CI: 2.92-4.84; p<0.001). Female children are more likely to be stunted compared to the male children (aOR= 1.55 95%CI: 1.37-1.74; p<0.001).

Children (0–59 months) who were not being breastfed at the time of the survey were more likely to be stunted compared to those who reported being breastfed at the time of the survey (aOR=1.20 95%CI: 1.04-1.38; p=0.015). Children whose birth weight was small or average were (aOR= 2.52 95%CI: 2.01-3.16; p<0.001 and (aOR= 1.43 95%CI: 1.24-1.65; p<0.001) 52% and 43% more likely to be stunted compared with children whose birth weight was large at birth.

## Discussion

The main objective of this study was to determine factors associated with stunting among children aged 0–59 months in Zambia during the period of 2018-2019. Bio-demographic and socio-economic factors contribute significantly to stunting in children. This study reveals that, stunting among children 0–59 months of age is still high in Zambia (34.9%) with Northern Province recorded the highest number of stunted children 45.5% followed by Luapula 44.8%.

Stunting increases as the age of a child (in months) increases; this is similar to what [4, Ghana] [3, Zambia] [11, Zambia] and [14, Ethiopia] found in their studies. This increase in stunting as age increases could be due to extended periods of inadequate food intake and increased morbidity among children 12–59 months.

Sex of a child and birth size showed significant association with stunting. Female children had higher odds of stunting than male, this was congruent with the studies conducted elsewhere [3, 5, 14].

Child birth size has a direct relationship with stunting. Small or average birth weight children were likely to be stunted compared with children whose birth weight was recorded as large; this is similar to what [14, Ethiopia] [8, Pakistan]. This could be linked to mother’s health and nutritional status before and during pregnancy which regulate the size of the child during intrauterine period as low birth weight is considered to be an indicator of restricted intrauterine growth.

This study also found that, children (0–59 months) who were not being breastfed at the time of the survey were more likely to be stunted compared to those who reported being breastfed. This could be that breastfeeding promotes child survival, health, brain development [2, 8]. Early initiation of breastfeeding prevents neonatal and infant deaths by reducing the risk of infectious diseases.

Maternal education play a significant role in determining stunting in children. Studies confirm that higher educated mothers understand and are act more responsively to the nutrition of their children, seek disease prevention and treatment, and maintain sanitary living. This could be because of exposure to media; they are likely to have better child and healthcare knowledge of nutrition. This study shows that children whose mothers had primary education were likely to be stunted compared to those whose mothers had secondary education. The same has been confirmed in previous studies and the results imply that maternal education may provide protective effects against all under-nutrition indicators in children [3, 5, 8, 14].

The study indicates that children from richer households were less likely to be stunted compared to those in poorer households. These findings are consistent with the other studies done on stunting [3, 5, 8,14]. This could be that that richer household are able to afford beyond basic needs such piped water, education, dietary foods, education, employment, and healthcare which considerably impact the health of their children. Children whose source of drinking water was non-improved were likely to be stunted compared to children whose source of water was improved. This may be attributed to the fact that non-improved water sources may be contaminated and thus may upsurge the risk of infection such as diarrhea. The study findings are consistent with other studies [11, 15]

## 5. Conclusions

Stunting is complex and influenced by multiple factors and structural determinants such as gender, age, water source, wealth quantile, birth weight, maternal education contribute to the reality that stunting is complex. These factors that influence stunting can be nutritional, socio-economic, demographic and environmental factors that impact on children’s health and wellbeing. These factors often not only influence stunting but also influence nutritional status of a child. Therefore, improvements in maternal education, household wealth, water and sanitation could improve health status of children. The findings of this study gives a benchmark of information on which planners and policy makers at different administrative levels should ride in formulating policies and programs aimed at improving the nutrition status of children under the age of 5 in Zambia.

## Data Availability

Dataset is available to third parties upon reasonable request to the corresponding author.

## Author Contributions

BN conceptualised the work, Data acquisition, analysis and interpretation. WFN and PJC supervised the analysis and interpretation. BN, WFN, PJC and ZM contributed critically to the revision of manuscript and approval of final version and accountable for all aspects of the work regarding its accuracy or integrity.

## Institutional Review Board Statement

Institutional authorization was obtained from University of Lusaka, School of Medicine & Health Sciences Research Ethics Committee (Reference: IORG0010092/ MPH19113810). National Health Research Authority (NHRA) granted permission (Ref No: NHRA00007/09/01/2021) in line with Act of Parliament (No. 2 of 2013) which mandatory requires all researchers to submit their research protocols to the NHRA upon receipt of approval from a Research Ethics Committee or an Institutional Review Board. No formal ethical clearance was obtained from ZSA as the study involved analysis of publically available data.

## Informed Consent Statement

Not applicable.

## Acknowledgments

This article is a part of the Master’s thesis submitted to the University of Lusaka in partial fulfillment of the requirements for the degree of Master of Public Health.

Glory be unto God for cherishing me through the entire period of learning and writing. Dr. Peter J. Chipimo (supervisor) for providing supervision throughout the research project. Mr. Wingston Ng’ambi of University of Malawi, nurturing me during this whole research project. Mr. Mowa Zambwe for organising all materials during article submission.

